# One-Sided Matching Portal (OSMP): a tool to facilitate rare disease patient matchmaking

**DOI:** 10.1101/2024.09.03.24313012

**Authors:** Matthew Osmond, E. Magda Price, Orion J. Buske, Mackenzie Frew, Madeline Couse, Taila Hartley, Conor Klamann, Hannah G. B. H. Le, Jenny Xu, Delvin So, Anjali Jain, Kevin Lu, Kevin Mo, Hannah Wyllie, Erika Wall, Hannah G. Driver, Warren A. Cheung, Ana S.A. Cohen, Emily G. Farrow, Isabelle Thiffault, Care4Rare Canada Consortium, Andrei L. Turinsky, Tomi Pastinen, Michael Brudno, Kym M. Boycott

## Abstract

**Background:** Genomic matchmaking - the process of identifying multiple individuals with overlapping phenotypes and rare variants in the same gene - is an important tool facilitating gene discoveries for unsolved rare genetic disease (RGD) patients. Current approaches are two-sided, meaning both patients being matched must have the same candidate gene flagged. This limits the number of unsolved RGD patients eligible for matchmaking. A one-sided approach to matchmaking, in which a gene of interest is queried directly in the genome-wide sequencing data of RGD patients, would make matchmaking possible for previously undiscoverable individuals. However, platforms and workflows for this approach have not been well established.

**Results:** We released a beta version of the One-Sided Matching Portal (OSMP), a platform capable of performing one-sided matchmaking queries across thousands of participants stored in genomic databases. The OSMP returns variant-level and participant-level information on each variant occurrence (VO) identified in a queried gene and displays this information through a customizable data table. A workflow for one-sided matchmaking was developed so that researchers could effectively prioritize the many VOs returned from a given query. This workflow was then tested through pilot studies where two sets of genes were queried in over 2,500 individuals: 130 genes that were newly associated with disease in OMIM, and 178 candidate genes that were not yet associated with a described disease-gene association in OMIM. These pilots both returned a large number of initial VOs (12,872 and 20,308, respectively), however the workflow successfully filtered out over 99.8% of these VOs before they were sent for review by a patient’s clinician. Filters on participant-level information, such as variant zygosity, participant phenotype, and whether a variant was also present in unaffected participants were especially effective in this workflow at reducing the number of false positive matches.

**Conclusions:** As demonstrated through the two pilot studies, one-sided matchmaking queries can be efficiently performed using the OSMP. The availability of variant-level and participant-level data is key to ensuring this approach is practical for researchers. In the future, the OSMP will be connected to additional RD databases to increase the accessibility of matchmaking to unsolved RGD patients.

## Background

In the past decade, genome-wide sequencing (GWS), including both exome sequencing and genome sequencing, has become a standard tool for the diagnosis of rare genetic diseases (RGDs) in many regions of the world. Depending on the indication(s) and specific implementation, it is estimated that approximately 40% of RGD families tested by GWS will receive a diagnosis.^1^ For comparison, the first-tier chromosomal microarray (CMA) test has an average diagnostic yield of 12.2% across a range of indications including developmental delay, dysmorphic features, intellectual disability, learning disabilities, autism and multiple congenital anomalies.^2^ And yet despite the success of clinical GWS testing, a significant portion of families with RGD remain undiagnosed following their clinical testing.

Families without a diagnosis following clinical GWS testing fall broadly into three categories: 1) the etiology of their condition is not monogenic, and unlikely to be resolved by current genetic testing methods; 2) the genetic etiology of their condition is not detectable by the GWS method employed, e.g. a complex structural variant with exome sequencing, but might be resolved by another technology; and 3) the genetic etiology of their condition is detectable by GWS, but the gene, region, or variant cannot yet be associated with disease, e.g., due to analytical challenges or insufficient evidence. In other words, families in this final group have a diagnosis within their existing sequencing data, but it is not recognized at the time of clinical testing.

Given the rapid pace at which new disease-gene associations and disease-variant associations are discovered each year, there is great interest in revisiting clinically generated GWS data to search for undetected molecular causes of RGDs. Many large-scale RGD research programs like the Undiagnosed Disease Network (USA, https://undiagnosed.hms.harvard.edu/), Genomics Answers for Kids (USA, https://www.childrensmercy.org/childrens-mercy-research-institute/studies-and-trials/genomic-answers-for-kids/), RD-Connect (EU, https://rd-connect.eu/) and our program discussed here, Care4Rare (Canada, https://www.care4rare.ca/), leverage this approach – conducting “reanalysis” of clinically generated GWS data to search for undetected molecular causes of RGD. A review of publications involving reanalysis of clinical GWS found the median new diagnostic rate was 15%, but that it varied considerably between studies.^3^ Working with GWS data in a research context broadens the tools and approaches that can be used to identify the molecular cause of the RGD. We recently performed a “clinical reanalysis” of a cohort of 287 families undiagnosed following clinical GWS.^4^ The reanalysis, limited to variants in genes known to be associated with disease in the Online Mendelian Inheritance in Man (OMIM) database, led to the identification of compelling candidate variants in 39 families (14%), and ultimately resulted in diagnoses for 13 families (5%). The most common factor in making these new diagnoses was the availability of new genomic knowledge, including new disease-gene associations, disease-variant associations, and phenotype expansions of existing conditions. These findings indicate that some undiagnosed RGD families will receive a diagnosis due to advances in global genomics knowledge if their data is revisited over time.

While periodic clinical reanalysis may capture diagnoses in newly described disease associations, it fails to address facilitating the discovery of the estimated thousands of novel disease-genes in real-time.^5^ RGD families with conditions with an unknown molecular etiology will remain undiagnosed until enough evidence is generated to associate a causal gene with their specific disease. An important criterion to substantiate such an association is the identification of multiple unrelated probands with pathogenic variants in the same gene and overlapping phenotype such that they are believed to have the same undescribed disease.^6^ The process of identifying similar families in this way is broadly called genomic matchmaking. Since RGDs are, by definition, extremely rare, the discovery of new disease genes hinges on worldwide sharing of information about undiagnosed families. To do this, tools like the Matchmaker Exchange (MME) enable submitters (who might be researchers, clinicians or RGD families) from around the world to connect and discuss potential overlap of cases.^7^ The MME uses a two-sided approach to matchmaking, in which submitters will only be matched if they have the same candidate gene flagged for their respective families. Since its inception in 2015, the MME has been heavily used by both research and clinical rare disease communities to facilitate the discovery of hundreds of novel disease-gene associations.^7^ Care4Rare’s own experience with two-sided matchmaking through the MME reflect its utility; over just two years, Care4Rare matched on 194 novel candidate genes, resulting in 861 connections with other submitters, ultimately leading to collaborations for 23 (15%) of these genes.^8^

While two-sided matchmaking has been a fundamental tool for gene discovery, the accessibility of this approach is limited for several reasons. Firstly, this hypothesis-based matching requires that each family have a candidate gene flagged (and thus must have been analyzed/reviewed) before it can be submitted. Secondly, while most databases currently connected to the MME support the inclusion of participant-level information (e.g., specific variant, zygosity, detailed phenotype, inheritance) when submitting a candidate gene for matchmaking, in our experience, most matches lack this information at the outset. Additional details on potential matches therefore must be exchanged by email after the initial match is made, which is time consuming for users.^8^ The current two-sided matchmaking model thus limits sharable data to the small set of families who: i) have been (re)analyzed and ii) have a flagged candidate gene, and limits submitters to those with the resources to follow up on many potential matches by email. Given the amount of GWS data that is produced through clinical and research testing,^9,10^ there are undoubtedly many untapped RGD datasets not currently available for two-sided matchmaking, restricting the ability to identify novel genetic etiologies for undiagnosed families.

Additional approaches to genomic matchmaking, which aim to address some of the shortcomings identified in two-sided matchmaking, have been proposed.^7^ One-sided matchmaking, involves a single party submitting a query on a gene or variant of interest to a database of GWS data to identify undiagnosed participants with variants matching the initial query. While a network of RGD databases like the MME has not yet been established for one-sided matchmaking, multiple databases have designed their own approaches to this type of matchmaking. MyGene2 (https://mygene2.org/MyGene2/), Geno2MP (https://geno2mp.gs.washington.edu/), VariantMatcher (https://variantmatcher.org/), and Franklin (https://franklin.genoox.com/) have designed variant-level implementations of one-sided matchmaking, in which users can search for the presence of a specific variant within the database, and receive phenotypic information on any participant found to carry this variant.^11^ Other platforms, such as the DatabasE of genomiC Variation and Phenotype in Humans using Ensembl Resources (DECIPHER),^12^ RD-Connect Genome-Phenome Analysis Platform (GPAP),^13^ and *seqr*^14^ support a gene-level approach to one-sided matchmaking instead, where a user can query a gene of interest and all variants in this gene are returned by the queried database. In evaluating these existing approaches to one-sided matchmaking, we see gene-level one-sided matchmaking as having the potential to identify variants of interest in undiagnosed RGD families through the querying of two types of genes: 1) genes that have been recently associated with human disease; and 2) genes that have not yet been associated with a disease. Given that a gene-level approach to one-sided matchmaking has the potential to, depending on the queried database’s size, return many variants, we expect additional data related to these variants will be crucial in ruling out false positive matches. To our knowledge, however, there has not been an assessment of what types of data and level of detail are needed to allow one-sided matchmaking users to filter gene-level queries down to a manageable number of variants of potential interest.

In this paper, we present a workflow for gene-level one-sided matchmaking and a beta version of a tool to support this approach, called the One-Sided Matching Portal (OSMP). The platform was designed with an emphasis towards providing a variety of participant-level and variant-level information inside a customizable interface, to best support users in making efficient one-sided matchmaking queries and limit the external communications required to rule potential matches in or out. To test the utility of the OSMP, as well as the one-sided matchmaking approach in general, we ran pilot studies using two sets of genes to identify new variants of interest in participants enrolled in Care4Rare: 1) 130 newly described OMIM disease genes - to search for disease-causing variants that would not have been recognized at the time of analysis by the clinical diagnostic laboratory; and 2) 178 novel candidate genes previously flagged on GWS analysis performed by the Care4Rare program.

## Methods

### The One-Sided Matchmaking Portal (OSMP)

We designed and built a tool, called the One-Sided Matching Portal (OSMP, https://github.com/ccmbioinfo/osmp), a web-based portal that can be connected to one or more RGD databases that contain variant and health information from research participants. The beta version of the OSMP supports gene-based queries of PhenoTips® instances by fetching and displaying single nucleotide variants and small insertions or deletions from the PhenoTips® variant store and participant information from PhenoTips® participant records.^15^ The OSMP’s frontend is written using the React JavaScript library (https://react.dev/), and the backend is designed using a Node.js framework (https://nodejs.org/). User authentication is managed using a Keycloak server (https://www.keycloak.org/), which supports a single sign-on for users using credentials from connected RGD databases.

A matchmaking query using the OSMP starts with the user defining a gene of interest, the maximum allele frequency of the returned variants (with a maximum value of 0.05), the RGD database(s) to be queried, and genome reference build in which to display the results. The OSMP sends the specified query to the selected database(s) and returns a table of variant occurrences (VOs, defined as a given variant in a given participant) meeting the specified criteria. The University of California Santa Cruz (UCSC) LiftOver tool (https://genome.ucsc.edu/cgi-bin/hgLiftOver) converts genomic coordinates of returned VOs to the user’s specified genome reference build, if necessary.

Each row of the result table corresponds to a specific VO, however, the rows can also be consolidated to display one unique variant per row. The results table is comprised of three categories of information returned for each VO, with multiple contributing data sources. The first category, variant information (Fig 1a), describes each variant, including its chromosome, genomic coordinates, reference allele, and alternate allele. This information is obtained from the PhenoTips® variant store, an indexed database sourced from variant files for each participant in the PhenoTips® instance. The second category, variant annotations (Fig 1b), involves more detailed variant information including predicted changes to the cDNA and amino acid sequences, the allele frequency in the gnomAD control database, and predictions of pathogenicity using the in-silico algorithms from Combined Annotation Dependent Depletion (CADD)^16^ and SpliceAI^17^. OSMP annotates this information at the time the query is returned using recent gnomAD^18^ and CADD^16^ annotations. This “on the fly” annotation was important to harmonize variant-level annotations within and between databases to accommodate the querying of source GWS files that may have been processed using different bioinformatic pipelines or at different times. This annotation step is currently performed for genes less than 200,000 bps in size due to computing limitations of the beta version of this platform. The final category of information, participant-level details (Fig 1c), includes information specific to the participant in whom each VO is found, including the zygosity of the variant, the clinical features in the form of a standardized vocabulary known as the Human Phenotype Ontology (HPO),^19^ and previously identified candidate genes. This information is extracted from the participant’s PhenoTips® record for each queried RGD database. The OSMP calculates some information on the fly including the number of heterozygous and homozygous participants returned for each VO (across all queried database(s)), as well as the number of VOs a participant has across the query gene (termed “burden”). Burden is especially important for identifying participants with potentially compound heterozygous VOs. A more detailed description of each data column returned or calculated by the OSMP is available in Additional File 1.

**Fig. 1.**
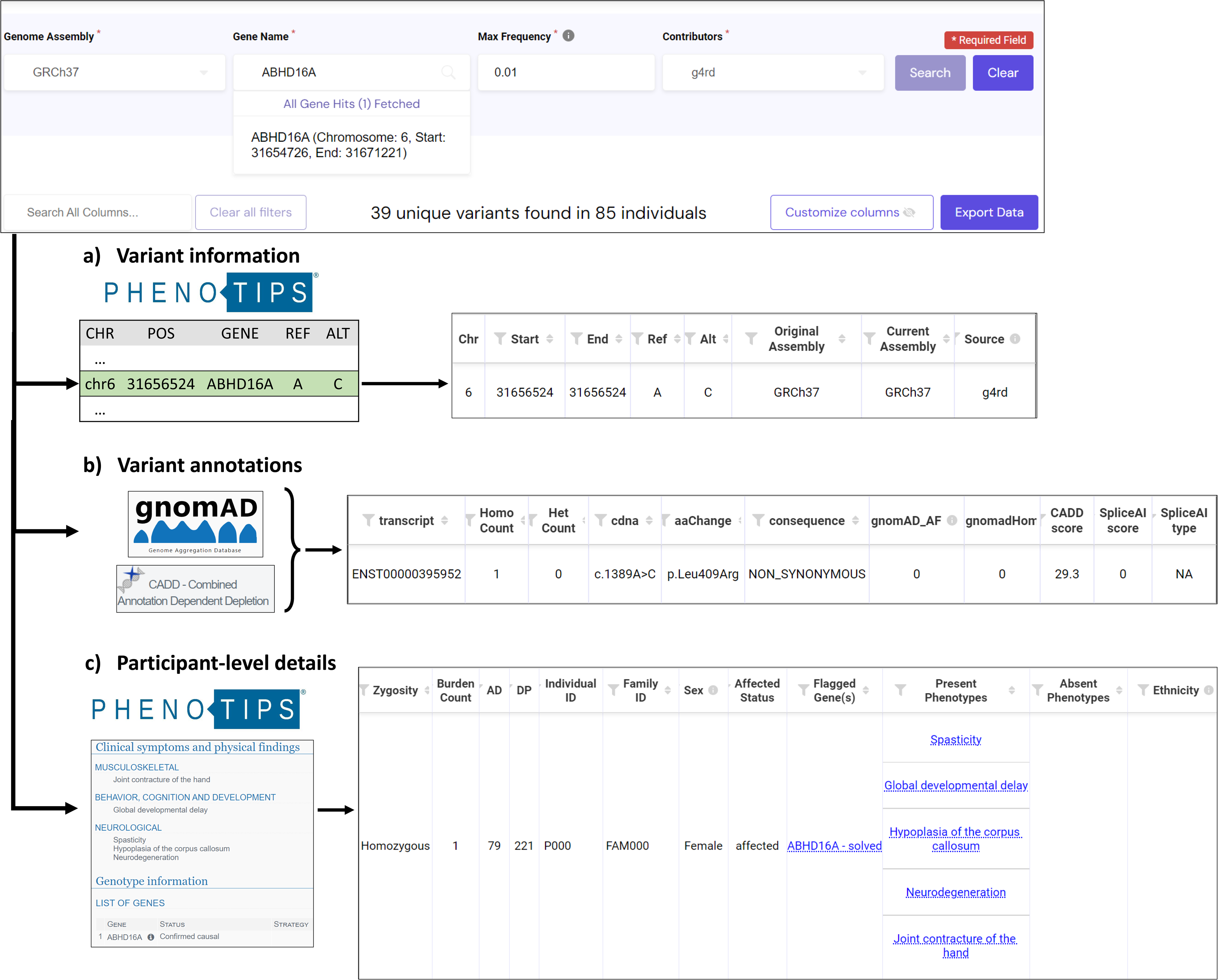
The One-Sided Matchmaking Platform (OSMP). Queries made for a gene of interest return data from three sources: **(a)** Variant information - basic information on variants identified in the PhenoTips® variant store. **(b)** Variant annotations - extracted from CADD and gnomAD datasets. **(c)** Participant-level details - phenotypic and genotypic details from individual PhenoTips® record.

The results table displayed by the OSMP is flexible, allowing users to customize the interface to best fit their workflow. Columns can be rearranged within the table or hidden, and each column can be filtered independently to narrow down to a list of VOs of interest. These filters are applied through the user’s local internet browser, meaning they are instantaneous and do not require an OSMP query to be rerun if filters are changed.

### Participant population

The beta version of the OSMP is connected to a single database, Genomics4RD,^20^ that houses data from thousands of participants enrolled in the Care4Rare Canada RGD gene discovery research program.^21^ Table 1 provides an overview of the participant demographics within Genomics4RD at the time of each of the two OSMP pilot studies. These participants include individuals who are affected with a RGD, many of whom are undiagnosed, as well as their unaffected family members. Affected participants in Genomics4RD are phenotyped using HPO terms, with participants having between 1 and 79 terms listed. Many of these affected participants presented with neurodevelopmental disorders, with the most common HPO terms across the database being global developmental delay (HP:0001263), seizures (HP:0001250), delayed speech and language development (HP:0000750), short stature (HP:0004322), and generalized hypotonia (HP:0001290). Numerous participants had one or more genes flagged in their record, classified as either as candidates, i.e., their role in the participant’s condition was inconclusive, or causal, i.e., they were a likely explanation for some or all of the participant’s presentation. Finally, participants had GWS data uploaded to their records, which had been aligned and annotated using the Care4Rare bioinformatics pipeline.^22^

**Table 1.**
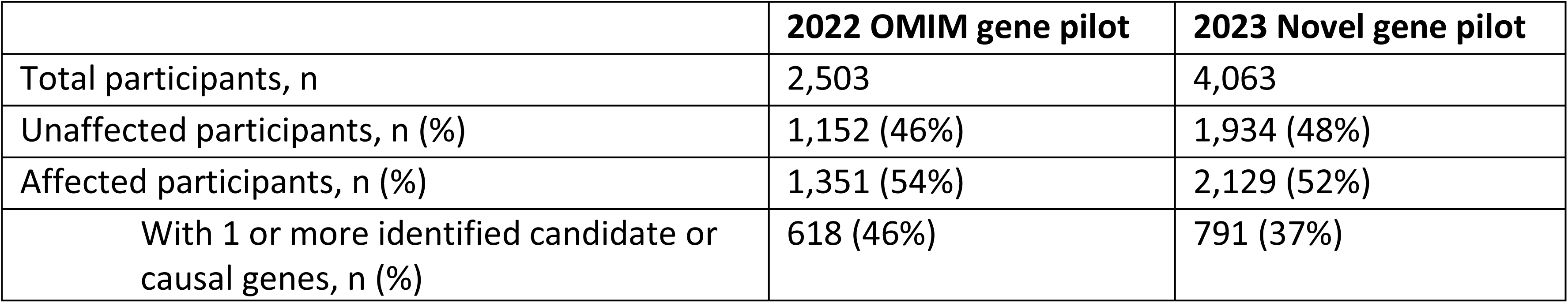
Genomics4RD participants at the time of each pilot study.

### A workflow for one-sided matchmaking

One-sided matchmaking on a gene level is expected to result in many potential matches, since it will return all VOs that exist for a given gene. We therefore devised a workflow to filter the large number of returned VOs down to a smaller, more manageable list of those most likely to be disease-causing (Fig 2). This seven-step workflow aimed to prioritize VOs that would be most likely to impact protein function and fit the inheritance pattern of a queried gene in participants with phenotypic overlap with the condition of interest.

**Fig. 2.**
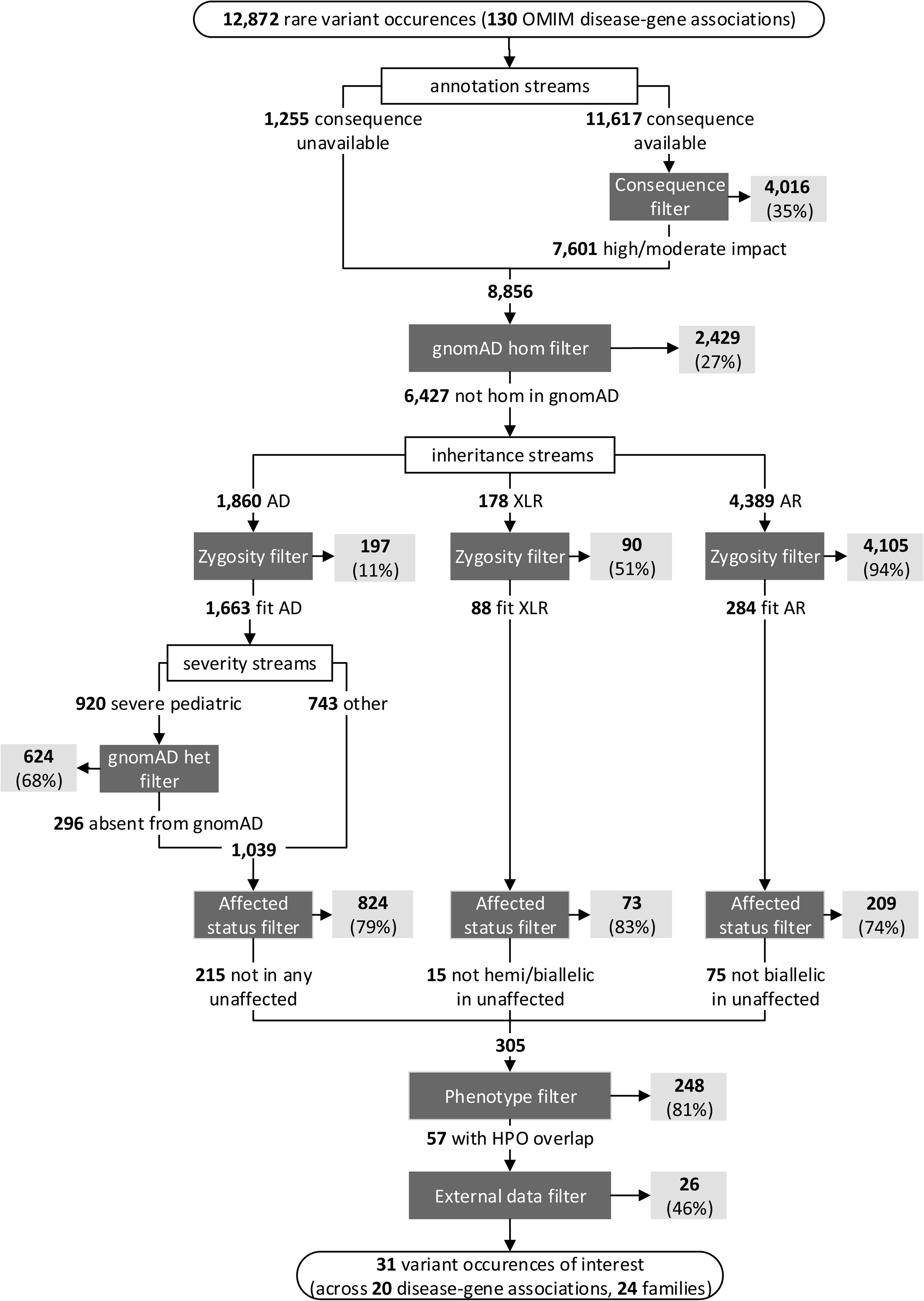
One-sided matchmaking workflow and details of variant occurrences (VOs) returned by the OMIM gene pilot queries (n=130 disease-gene associations). Filtration steps are indicated by dark grey boxes, and VOs removed by each filter is indicated by light grey boxes. Percentages indicate the proportion of VOs from the previous step that were filtered out. AD: autosomal dominant, XLR: X-linked recessive, AR: autosomal recessive.

The first applied filter was based on predicted protein function (i.e., the **consequence filter**). VOs that were predicted by Ensembl’s Variant Effect Predictor (VEP)^23^ to have a low impact on protein behaviour (i.e., variants located outside of any exon or splice site) and had a SpliceAI score less than 0.5 were filtered out. VOs that were not given a conclusive annotation by VEP bypassed this filter and proceeded to the next step.

Next, VOs seen at least once in a homozygous state in gnomAD were removed (i.e., **gnomAD hom filter**), as homozygous variants present in reportedly unaffected participants are unlikely to be disease-causing, regardless of the inheritance pattern for a disease-gene association.

At the third stage of the workflow, we defined three streams based on different inheritance patterns for a given disease-gene association: an autosomal dominant (AD) stream, an X-linked recessive (XLR) stream, and an autosomal recessive (AR) stream. In each stream, a **zygosity filter** was first applied, meaning that only heterozygous VOs were kept in the AD stream, hemizygous VOs were kept in the XLR stream and homozygous, or occurrences of multiple heterozygous VOs in the same participant, were kept in the AR stream. For VOs in genes associated with AD disease with a severe pediatric onset, we applied an additional filter to remove variant occurrences present in any zygosity in gnomAD (i.e., **gnomAD het filter**), since this control database is expected to be largely absent of severe pediatric disease.^18^

Next, we used the queried genomic database as a control cohort, similar to how the gnomAD filters were used. If a VO was seen in an unaffected participant with the inheritance pattern anticipated for the disease-gene association, the VO was filtered out (i.e., **affected status filter**). For example, when querying an AR disease-gene association, a homozygous VO in an unaffected participant would result in the VO being filtered out for them and any other participant homozygous for that same variant.

Finally, we were left with a prioritized list of VOs with which to perform phenotype/genotype correlation with the original disease-gene query. The HPO terms of each participant with a remaining VO were compared to the clinical descriptions of the disease being queried by a certified genetic counsellor (Author MO). VOs in participants with insufficient phenotype overlap with queried disease features were removed (i.e., **phenotype filter**), and remaining VOs were then manually reviewed using external information sources (i.e., **external data filter**). Such information included ClinVar variant classifications, detailed clinical notes, and sequencing data from family members not included in the RGD database. The VOs remaining following this final filter were reviewed by a multidisciplinary team including medical geneticists, laboratory geneticists, and genetic counsellors to determine if they warranted clinical validation or further investigation.

## Results

### One-Sided Matchmaking Pilot – OMIM Genes

Our first pilot of the one-sided matchmaking workflow using the beta version of the OSMP involved querying a set of genes recently associated with disease in OMIM (i.e., OMIM gene pilot). As these disease-gene associations would not have been documented at the time of clinical GWS analysis, we hypothesized this gene set may be enriched for disease-causing variants in unsolved RGD patients in the Genomics4RD database. First, a list of new disease-gene associations added to OMIM between November 2021 and August 2022 was generated (n=227). These associations were then narrowed down to genes with sizes that fit within the OSMP’s current memory limitation of 200,000bps. Finally, the remaining associations were manually reviewed to prioritize diseases with presentations most relevant to the Genomics4RD patient population (i.e., diseases that severely impact a single system, or multiple systems), resulting in a final list of 130 disease-gene associations (across 116 unique genes, i.e., 14 genes had two newly described disease associations) to be queried using the OSMP. Approximately 63% (82/130) of the disease-gene associations were for AR conditions, 32% (42/130) for AD conditions, and 5% (6/130) for XLR conditions.

In total, 12,872 VOs with an allele frequency of 0.01 or less in Genomics4RD were returned by the OSMP across the 130 disease-gene associations queried. Figure 2 and Table 1 detail the number and proportions of VOs removed at each stage of the workflow, respectively. The consequence filter removed 35% of the VOs with a VEP-annotated impact. Next, filtering out VOs seen in a homozygous state in gnomAD removed 27% of remaining VOs. The effectiveness of the zygosity filter varied across the inheritance patterns, with 11% of the remaining VOs filtered out VOs for AD associations, 51% of VOs for XLR associations, and 94% of VOs for AR disease-gene associations. For early-onset AD conditions, 68% of the VOs that passed the zygosity filter were removed due to their presence in the gnomAD database. An additional 79% of the VOs for AD disease-gene associations, 83% for XLR associations, and 74% of VOs for AR associations were removed as they were present in the anticipated zygosity in unaffected participants. This resulted in 305 VOs remaining after all filters using OSMP-provided data. Across all inheritance patterns, 81% of the remaining VOs were filtered out due to insufficient phenotypic overlap with the disease synopsis in OMIM. Finally, 46% of the remaining VOs were removed following a manual review using data sources external to the OSMP. We filtered out VOs at this stage due to ClinVar classifications as benign or likely benign, variants not segregating appropriately in family members, and insufficient phenotypic overlap with the OMIM disease following review of external clinical notes. Following all filtration steps, a total of 31 VOs (0.24% of the original query results) remained across 20 newly described disease-gene associations and were prioritized for review with the multidisciplinary team: 70% (14/20) of these disease-gene associations were for autosomal dominant conditions, and 30% (6/20) were for autosomal recessive conditions. Of these, one, so far, has resulted in a diagnosis. A de novo VO in the gene *POLR3B* was identified in a patient with overlapping neurological features to the recently described association with demyelinating Charcot-Marie-Tooth disease type 1I (OMIM 619742). This variant, identified through the OMIM gene pilot, has since been classified as likely pathogenic through validation by a clinical diagnostic laboratory.

### One-Sided Matchmaking Pilot – Novel Genes

The second one-sided matchmaking pilot queried a set of candidate genes assigned to a disease-gene association in OMIM (designated as the novel gene pilot). These candidate genes were identified through the analysis of GWS data for unsolved RGD patients enrolled in the Care4Rare research program (criteria described by Osmond et al).^8^ Most of these participants have records in the Genomics4RD database and are queriable via the OSMP. This second pilot had two goals: first, to validate the one-sided matchmaking workflow (i.e., do we identify the true positive participants who harbor compelling candidates in these genes), and second to identify additional families with rare variants in the same gene and overlapping phenotype that would help to build evidence for a novel disease-gene association. A list of 178 novel candidate genes was queried using the OSMP for this novel gene pilot. Approximately 62% (111/178) of these genes were hypothesized to be associated with an AD condition, 33% (58/178) were hypothesized to be associated with an AR condition, and 5% (9/178) were hypothesized to be associated with an XLR condition. Of these 178 candidate genes, 140 had true positive participants with data accessible to the OSMP.

### Validation of the one-sided matchmaking workflow

We queried OSMP with the set of 140 genes with VOs previously prioritized as disease-causing or strong candidates in Care4Rare families to test the one-side matchmaking workflow. After applying all filters, the VOs from 89% (124/140) of these genes remained within our prioritized list. For the 16 genes with VOs that did not pass all filters, six genes had VOs that were removed by the **consequence filter**, most commonly because the VOs occurred just outside a canonical splice site. Nine had VOs removed by the **gnomAD het filter**, as these genes were associated with a severe AD pediatric onset condition, but the VO was present in at least one individual in gnomAD. In all these cases, the presence of the variant in gnomAD had been previously identified and the variant was considered a weak novel candidate. Finally, one gene had a VO that was removed by the **affected status filter**, as the variant was present in a family member marked as ‘unaffected’, however on reflection it was deemed that the affected status of this relative was inconclusive. Upon review, we did not believe that these false negatives warranted changes to our one-sided matchmaking protocol.

### Use of OSMP to identify additional novel candidate gene families

We excluded the true positive families described above in our candidate gene query of OSMP for additional families with seemingly the same novel RGD. In total, 20,308 VOs with a maximum allele frequency of 0.01 in Genomics4RD were returned by the OSMP related to the 178 candidate genes. Figure 3 and Table 2 detail the number and proportions of VOs removed at each stage of the workflow, respectively. Overall, the **consequence filter** removed 28% of eligible VOs, and the **gnomAD het filter** removed 25% of the remaining VOs. Like the OMIM gene pilot, the efficacy of the **zygosity filter** differed between the hypothesized inheritance patterns for these novel genes. Approximately 3% of VOs for AD genes, 65% of VOs for XLR genes and 91% of VOs for AR genes were removed by this filter. Filtering out variants seen in gnomAD removed 75% of the remaining VOs from genes with a suspected early-onset AD condition. When filtering out VOs seen in unaffected participants, 82% of the VOs for AD disease-gene associations, 45% for XLR associations and 87% for AR associations were removed. This resulted in 604 VOs remaining after all filters using OSMP-provided data. Filtering out participants with insufficient phenotype overlap to the original patient in which the novel gene was identified resulted in the removal of 91% of VOs across all patterns of inheritance. Finally, approximately 67% of the remaining VOs were removed using data not available directly through the OSMP. External data used to rule out these VOs was similar to the OMIM gene pilot and included sequencing data from family members not in Genomics4RD, more extensive notes on clinical presentation, and the number of hemizygotes who carry a variant in gnomAD for XLR genes. Following all filtration steps, a total of 18 VOs (0.09% of the initial query results) across 14 novel candidate genes remained for review with the multidisciplinary team. Ten of these novel candidate genes were hypothesized to be associated with AD conditions (10 heterozygous VOs), three genes were hypothesized to be associated with AR conditions (2 homozygous VOs, and 2 heterozygous VOs in the same participant), and one gene was thought to be associated with a XL condition (1 hemizygous VO). One of these remaining VOs is in the gene *CCDC6* and it is believed to be the molecular cause for this participant’s rare disease. This heterozygous *CCDC6* variant has already been described in several other individuals in an ongoing collaboration first established through the MME, and this participant represents an addition to this cohort. Review of the other prioritized VOs is ongoing.

**Fig 3.**
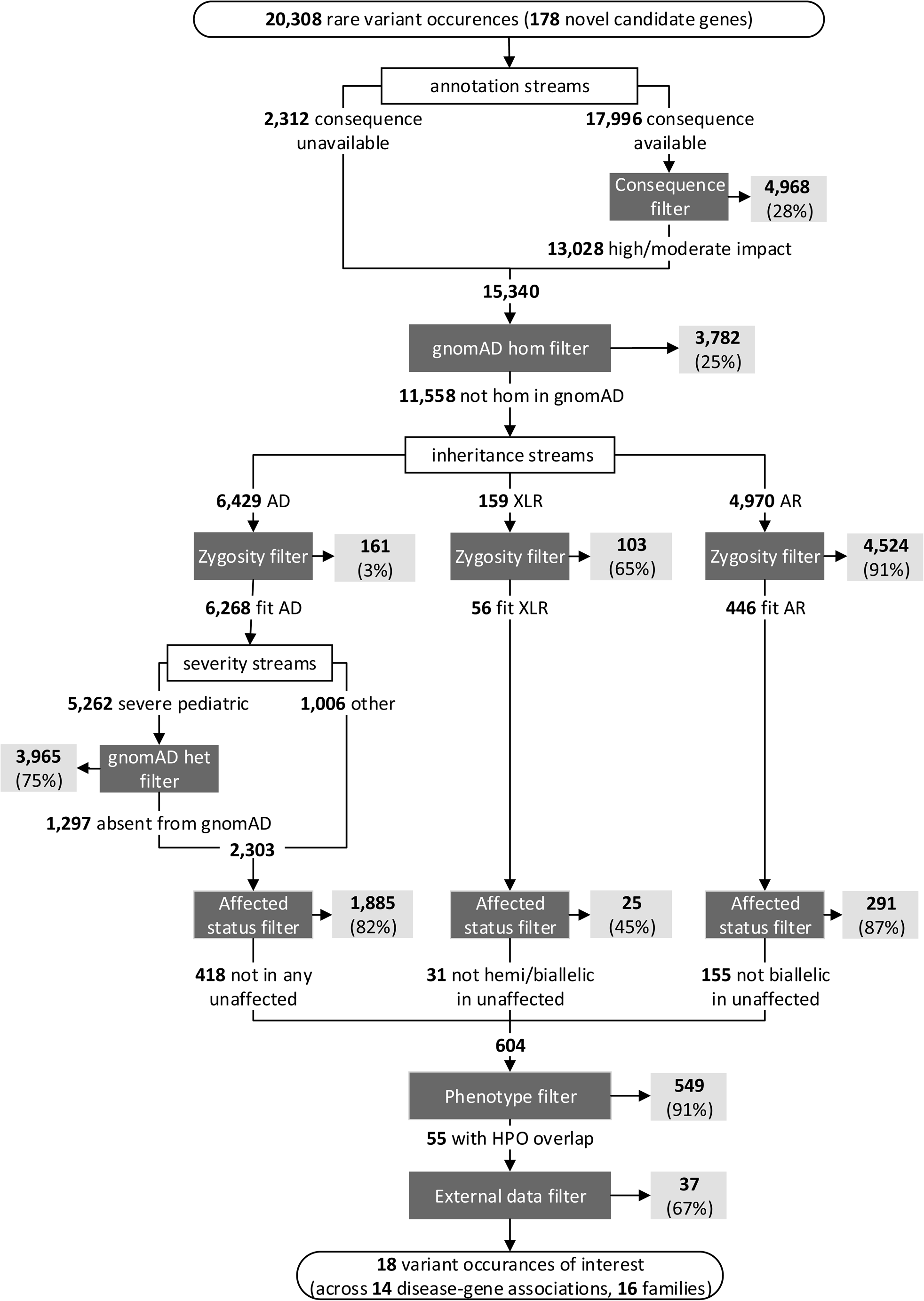
One-sided matchmaking workflow for variant occurrences (VOs) returned by the novel gene pilot queries (n = 178 novel candidate genes). Filtration steps are indicated by dark grey boxes, and VOs removed by filters are indicated by light grey boxes. Percentages indicate the proportion of VOs from the previous step that were filtered out. AD: autosomal dominant, XLR: X-linked recessive, AR: autosomal recessive.

**Table 2.**
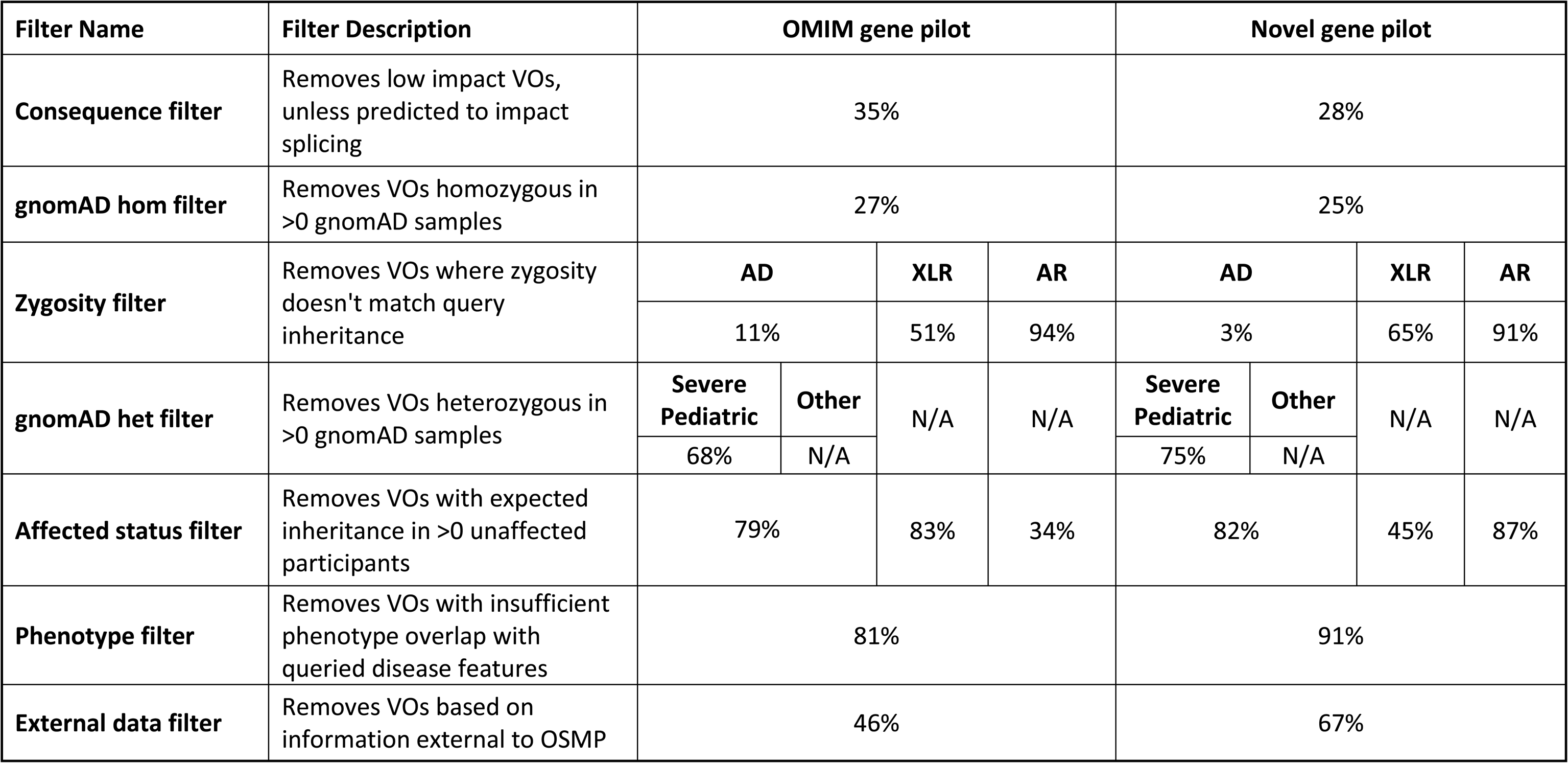
Efficacy of workflow filters in the OMIM gene and novel gene pilots. Percentages indicate the proportion of remaining VOs removed by each filter step.

## Discussion

The development and piloting of the beta version of the OSMP shows that one-sided matchmaking can be effective in identifying genetic variants of interest in undiagnosed patients with RGD. Though tens of thousands of VOs were returned in each pilot, we developed an effective workflow to quickly filter VOs to those most likely to be disease-causing. While clinical review of the prioritized VOs is ongoing, there has been one diagnosis made in the newly described OMIM gene *POLR3B*, and one diagnosis made in the novel gene *CCDC6* where collaboration is ongoing. We anticipate that further review of prioritized VOs will lead to diagnoses in additional genes.

The OMIM gene and novel gene pilots highlight the importance of making participant-level information available when performing this type of matchmaking to rule out as many false positives as possible before undergoing more extensive case reviews. Knowing when variants are present in unaffected participants was highly effective in filtering VOs across genes associated with all inheritance patterns, in total removing 78% and 79% of the remaining VOs in the OMIM gene and novel gene pilots, respectively. Similarly, access to phenotypes in the form of HPO terms for affected participants enabled the removal of over 80% of the remaining VOs across both one-sided matchmaking pilots. Lastly, zygosity information on VOs was especially effective as a filter for genes with a known or hypothesized AR inheritance pattern, resulting in the removal of over 90% of remaining VOs for both pilot studies. The utility of phenotypic and genotypic data in proactively ruling out potential matches is a trend that our team has also experienced with two-sided matchmaking - over half of two-sided matches were ruled out when such information was available at the time the initial match was made.^8^ Increasing the inclusion of such participant-level information for matches submitted to the Matchmaker Exchange has been highlighted as an important factor to improve the efficiency of two-sided matchmaking,^7^ and our pilots suggest that one-sided matchmaking platforms will not be successful without this data being made available.

While the beta version of the OSMP can provide the information necessary to make efficient one-sided matchmaking queries through an interface that is easy to use and customizable, these pilot studies provide insight into ways that the platform can be further improved for more widespread use. Providing on the fly annotations with CADD and gnomAD datasets ensures variant-level data returned by the OSMP remains accurate and harmonized across databases using different bioinformatics pipelines, however this feature is currently limited to genes less than 200,000 base pairs in size. Allocation of additional computing resources or performance improvements to the existing software may be required to ensure this feature is available to queries of genes of all sizes. Reviewing the VOs that were ruled out using data not directly available through the OSMP also indicates ways in which the platform can be improved. Knowledge of whether a variant has been classified in ClinVar, or if variants in XLR genes are seen in a hemizygous state in gnomAD, would both be useful to have available in the OSMP annotations. More detailed information on the inheritance of VOs (i.e., if a heterozygous variant is de novo vs inherited, or if multiple heterozygous variants are in cis or trans) would also improve the efficiency of one-sided matchmaking on this platform, as the results generated by the OSMP are not currently optimized for family-based analyses.

Finally, future versions of the OSMP will be expanded in both the number of users with access to the platform, and the number of connected databases. Upcoming OSMP development will focus on load testing the tool so that more users – both members of the Genomics4RD database and other third-party researchers - can utilize this resource. In this next phase, connecting additional databases to the OSMP will be crucial for both increasing the number of unsolved RGD patients made available for one-sided matchmaking and improving the OSMP’s ability to rule out existing VOs of interest with an increased number of internal controls (i.e., unaffected family members). The OSMP is in the process of establishing a connection to the PhenoTips® database maintained by the Genomic Answers for Kids rare disease research program, which will enable the OSMP to query over 2,900 additional RGD participants with GWS data.

### Conclusions

The beta version of the OSMP can perform gene-level one-sided matchmaking queries for the purposes of prioritizing variants of interest in undiagnosed RGD patients. The development and piloting of the one-sided matchmaking workflow for both newly described disease-gene associations and novel candidate genes demonstrates both the sheer number of variant occurrences returned by gene-level queries, and the importance of variant-level and participant-level data in filtering possible matches down to a level more reasonable for users to review. Further, our pilots act as proof-of-principle that one-sided matchmaking can identify additional diagnoses and candidate genes. The lessons learned from piloting the beta version of the OSMP will be used to further refine the functionality of the platform, and we believe these insights will be of use to other groups developing similar tools. The connection of additional RGD databases to one-sided matchmaking services like the OSMP will be crucial in providing access to matchmaking for as many clinically undiagnosed RGD families as possible, in the hopes of identifying the genetic etiologies of their conditions.

## Declarations

### Ethics approval and consent to participate

Informed consent was obtained for all participants that were queried by the OSMP in this study. Participant-level data made available for the purposes of matchmaking was de-identified. Institutional review board approval was obtained from the Children’s Hospital of Eastern Ontario for both Finding of Rare Disease Genes in Canada (FORGE) and Care4Rare Canada (Research Ethics Board # 11/04E). Institutional review board approval for Care4Rare-SOLVE was obtained from Clinical Trials Ontario (CTO1577).

## Consent for publication

Not applicable.

## Availability of data and materials

The source code for the OSMP is available at https://github.com/ccmbioinfo/osmp. Access to the beta version of the OSMP is currently limited to a small set of Genomics4RD users. The genotypic and phenotypic data that support the findings of this study are located in the controlled access database Genomics4RD. Genomics4RD open access data is available at https://www.genomics4rd.ca/.

## Competing interests

OJB and MB have an equity interest in, and OJB is an employee of PhenoTips®, which licenses software used in the Genomics4RD database.

## Funding

This study was funded through the *Genomics and Precision Health Top-up* grant GPT-174518 “Care4Rare-Solve: Efficient cross-border matchmaking to deliver diagnoses for rare genetic diseases”, awarded by the Canadian Institutes of Health Research.

The development of Genomics4RD and the production of its housed GWS data was performed under the Care4Rare Canada Consortium funded by Genome Canada and the Ontario Genomics Institute (OGI-147), the Canadian Institutes of Health Research, Ontario Research Fund, Genome Alberta, Genome British Columbia, Genome Quebec, and Children’s Hospital of Eastern Ontario Foundation.

T.H. was supported by a Frederick Banting and Charles Best Canada Graduate Scholarship Doctoral Award from Canadian Institutes of Health Research. MB is a CIFAR AI Chair. KMB was supported by a Canadian Institutes of Health Research Foundation grant FDN-154279 and a Tier 1 Canada Research Chair in Rare Disease Precision Health. The work at Children’s Mercy Kansas City is supported by generous donors to Children’s Mercy Research Institute and Genomic Answer For Kids program.

## Authors’ contributions

Funding acquisition for this study was conducted by EMP, TH, TP, MB, and KMB. MO, EMP, OJB, TH, CK, ALT, TP, MB, and KMB contributed to the conceptualization and high-level design of OSMP. Development and testing of OSMP was performed by MO, EMP, OJB, MF, MC, CK, HGBHL, JX, DS, KL, KM, HW, HGD, ALT, and MB. Curation of the data to be queried by OSMP was conducted by EMP, MC, AJ, HW, HGD, WC, AC, EF, and IT. MO, EMP, and TH designed the one-sided matchmaking workflow. MO, HW, and EW collected the data generated from the OSMP pilot studies. Data analysis from the pilot studies was performed by MO and EMP. The initial manuscript draft was written by MO, EMP, and KMB. All authors have read and approved the final manuscript.

## Supporting information

Supplemental file 1: Overview of data columns returned or generated by the OSMP

## Acknowledgements

We would like to acknowledge Alina Gvozdik, John Q. Miller, Joel John, Kevin M. Power, John N. Gregor, and Bourke B. Hutchison for their contributions towards establishing API connections between the OSMP and its connected databases.

## Additional Files

**Additional file 1.pdf** Overview of data columns returned or generated by the OSMP

## List of abbreviations

AD: Autosomal dominant
AR: Autosomal recessive
CADD: Combined Annotation Dependent Depletion
CMA: Chromosomal microarray
DECIPHER: DatabasE of genomiC varIation and Phenotype in Humans using Ensembl Resources
FORGE: Finding of Rare Disease Genes
GPAP: Genome-Phenome Analysis Platform
GWS: Genome wide sequencing
HPO: Human Phenotype Ontology
MME: Matchmaker Exchange
OMIM: Online Mendelian Inheritance in Man
OSMP: One-Sided Matching Portal
RGD: Rare genetic disease
UCSC: University of California Santa Cruz
VEP: Variant Effect Predictor
VO: Variant occurrence
XLR: X-linked recessive

## Notes

### Funding Statement

This study was funded through the Genomics and Precision Health Top-up grant GPT-174518 titled ″Care4Rare-Solve: Efficient cross-border matchmaking to deliver diagnoses for rare genetic diseases″, awarded by the Canadian Institutes of Health Research. The development of Genomics4RD and the production of its housed GWS data was performed under the Care4Rare Canada Consortium funded by Genome Canada and the Ontario Genomics Institute (OGI-458 147), the Canadian Institutes of Health Research, Ontario Research Fund, Genome Alberta, Genome British Columbia, Genome Quebec, and Children″s Hospital of Eastern Ontario Foundation. T.H. was supported by a Frederick Banting and Charles Best Canada Graduate Scholarship Doctoral Award from Canadian Institutes of Health Research. MB is a CIFAR AI Chair. KMB was supported by a Canadian Institutes of Health Research Foundation grant FDN-154279 and a Tier 1 Canada Research Chair in Rare Disease Precision Health. The work at Children″s Mercy Kansas City is supported by generous donors to Children″s Mercy Research Institute and Genomic Answer For Kids program.

### Author Declarations

The institutional review board of the Children″s Hospital of Eastern Ontario gave ethical approval for this work. The institutional review board of Clinical Trials Ontario gave ethical approval for this work.

