## Supplemental file 1: Overview of data columns returned or generated by the OSMP for "One-Sided Matching Portal (OSMP): a tool to facilitate rare disease patient matchmaking"

### Additional file 1 – Overview of data columns returned or generated by the OSMP

| Table Section | Column Name | Description | Source | Source Variable | Example |
|---|---|---|---|---|---|
| Variant information | Chr | Chromosome | Phenotips - Variant Store | chromosome | 6 |
| | Start | Starting position of the variant (based on current assembly) | Phenotips - Variant Store | start | 31656524 |
| | End | End position of the variant (based on current assembly) | Phenotips - Variant Store | end | 31656035 |
| | Ref | Reference allele. More than one nucleotide listed as the reference allele means there is a deletion at this position. | Phenotips - Variant Store | ref | A |
| | Alt | Alternative allele. More than one nucleotide listed as the alternative allele means there is a deletion at this position. | Phenotips - Variant Store | alt | G |
| | Current Assembly | The assembly used to annotate variants in the OSMP query results | OSMP - calculated from "Genome Assembly" search field | assemblyIdCurrent | GRCh37 |
| Variant details | transcript | We select the transcript that is most severely affected using the ConsScore as annotated in the CADD VCF; the ConsScore corresponds to the VEP severity listed in https://grch37.ensembl.org/info/genome/variation/prediction/predicted_data.html. If there is a tie, the transcript that appears first as listed by CADD is chosen. | CADD | FeatureID | ENST00000395952 |
| | Homo Count | Number of participants in the OSMP results who are homozygous for this variant. Note that this count is summed across all databases that were queried | OSMP - calculated | homozygousCount; zygosity has "hom" in it | 1 |
| | Het Count | Number of participants in the OSMP results who are heterozygous for this variant. Note that this count is summed across all databases that were queried | OSMP - calculated | heterozygousCount; zygosity has "het" in it | 14 |
| | cdna | Coding DNA change according to the Ensembl transcript. Note that this column will appear as "NA" for any variants that are not located in an exon. | CADD | cdna | c.1389A>C |
| | aaChange | Amino acid change according to the Ensembl transcript. Note that this column will appear as "NA" for any variants that are not located in an exon. | CADD | rAA | p.Leu409Arg |
| | consequence | Calculated variant consequence according to the Ensembl Sequence Ontology | CADD | Consequence | NON_SYNONYMOUS |
| | gnomAD_AF | Allele frequency of this variant in gnomAD. The AF reported is the HIGHEST population frequency between the exome AF and the genome AF. | gnomAD | Max of AF_popmax from gnomAD exome and gnomAD genome | 0.00131885 |
| | gnomAD_AC | Allele count of this variant in gnomAD. Note that unlike the gnomAD_AF column this is the SUM of the allele counts across the exome and genome data. | gnomAD | Sum of allele counts across gnomAD exome and gnomAD genome | 5 |
| | gnomadHom | Number of individuals in gnomAD homozygous for this variant. Note that unlike the gnomAD_AF column this is the SUM of the homozygotes across the exome and genome data. | gnomAD | sum of nhomalt from gnomAD exome and gnomAD genome | 2 |
| | CADD score | Scaled score for predicting the deleteriousness of single nucleotide variants, insertions, and deletions. A score of 20 or greater indicates a variant in the top 1% for likelihood of being deleterious. | CADD | PHRED | 24.3 |
| | SpliceAI score | SpliceAI probability of a variant being splice altering (scale of 0 to 1). Closer to 1 means a greater probability of a splicing change, recommended cutoff is normally 0.5. | CADD | Maximum of [SpliceAI-acc-gain, SpliceAI-acc-loss, SpliceAI-don-gain, SpliceAI-don-loss] | 0.09 |
| | SpliceAI type | SpliceAI type of splicing change predicted for this variant. Can be a gain or a loss of a splice acceptor or splice donor. If the SpliceAI score is 0, this will be "NA". | CADD | Splice change type associated with SpliceAI | SpliceAI-don-loss |

| | | | | | |
|---|---|---|---|---|---|
| | | | | score above. NA if 0 for all types. | |
| Participant-level details | Original Assembly | The assembly that variants were annotated with in the source database | Phenotips - Variant Store | assemblyId | GRCh38 |
| | Source | The source database where this participant is located. | OSMP | source | g4rd |
| | Zygosity | Zygosity for the variant in this participant. Note that hemizygous variants in this column will appear as "homozygous". | Phenotips - Variant Store | zygosity | Heterozygous |
| | Burden Count | Number of variants a participant has in this gene. Note that this is calculated AFTER variants have been returned by the search query, meaning that variants more common than the "Max Frequency" will NOT be included in the burden calculation. | OSMP - calculated from the number of variant in the displayed results | burdenCount | 1 |
| | AD | Allele depth. This is the number of reads in which a variant is called for a given sample. | Phenotips - Variant Store | ad | 79 |
| | QUAL | Phred-scaled probability that a variant exists given sequencing data. A phred score of 20 indicates that a variant is 99% accurate, with a 1% chance of error. | Phenotips - Variant Store | QUAL | 3929.2 |
| | Individual ID | Identifier for this participant in its source database | Phenotips - Patient Record | individualId | P0000001 |
| | Family ID | Identifier for this participant's family in its source database | Phenotips - Patient Record | familyId | FAM0000001 |
| | Sex | Biological sex of this participant | Phenotips - Patient Record | sex | Female |
| | Diseases | Diseases that have been clinically diagnosed in this participant. These are only diseases that have been listed as an "OMIM diagnosis" in Phenotips. | Phenotips - Patient Record | disorders | THREE M SYNDROME 2 (MIM:612921) |
| | Affected Status | Flag for whether a participant is clinically affected or unaffected. Note that participants are only listed as unaffected if "This patient is clinically normal" is checked off in Phenotips. | Phenotips - Patient Record | clinical-diagnosis | affected |
| | Flagged Gene(s) | Gene(s) explicitly flagged for this participant record, along with their classification. Note that genes flagged as "solved" may be only a partial explanation for an participant's phenotype. | Phenotips - Patient Record | genes + status | ABHD16A - solved |
| | Present Phenotypes | HPO phenotypes listed as present in this participant (click on text to expand field) | Phenotips - Patient Record | phenotypicFeatures and clinicalStatus | Spasticity<br>Global developmental Delay<br>Neurodegeneration |
| | Absent Phenotypes | HPO phenotypes explicitly listed as absent from this participant (click on text to expand field) | Phenotips - Patient Record | phenotypicFeatures and clinicalStatus | Premature birth<br>Abnormal delivery |
| | Ethnicity | Parental ethnicities for this participant. | Phenotips - Patient Record | ethnicity | French Canadian, Jamaican |
| | Contact | Contact information for the owner of the participant record | OSMP | contactInfo | Care4Rare Admin |
